# Data-driven personalised recommendations for eczema treatment using a Bayesian model of severity dynamics

**DOI:** 10.1101/2024.01.21.24301575

**Authors:** Guillem Hurault, Jean François Stalder, Markéta Saint Aroman, Reiko J. Tanaka

**Author notes:** **Corresponding author:** Reiko J. Tanaka. **Data accessibility:** The analysis code is available at https://github.ic.ac.uk/tanaka-group/EczemaTreat. **Authors’ contributions** GH: Conceptualization, data curation, formal analysis, investigation, methodology, software, validation, visualization, writing-original draft. JFS, MSA: Resources. RJT: Conceptualization, funding acquisition, project administration, resources, supervision, validation, writing-original draft, writing-review & editing.

## Abstract

Atopic dermatitis (AD) is a chronic inflammatory skin disease. AD has heterogeneous phenotypes, making it challenging to predict treatment effects for each patient and to generate personalised treatment recommendations. Here we aim to develop a computational model that predicts the evolution of AD severity and generates treatment recommendations for individual patients. We modelled the temporal evolution of eczema severity by applying a previously developed computational framework (EczemaPred) to the daily record of Patient-Oriented SCORing Atopic Dermatitis (PO-SCORAD) collected from 16 AD patients over 12 weeks in an observational study. We also leveraged historical data from 337 AD patients to kickstart the model training and reach more robust conclusions. We estimated the effects of topical corticosteroids and emollients on the next day’s PO-SCORAD, and generated personalised treatment recommendations using Bayesian decision analysis on whether treatment should be applied to improve PO-SCORAD on the next day for each of the 16 patients. We calibrated daily PO-SCORAD recorded by patients with monthly SCORAD assessed by clinical staff to improve the data quality. This study demonstrated a proof-of-concept for generating personalised treatment recommendations for AD using a Bayesian model that integrates multiple sources of information, including PO-SCORAD, SCORAD, and treatment usage.

## INTRODUCTION

Atopic dermatitis (AD or eczema) is a chronic inflammatory skin disease characterised by dry, itchy skin [1]. The current treatments mainly aim to manage disease symptoms using, e.g. topical corticosteroids and emollients. Given the heterogeneity of the disease symptoms and treatment responses, designing personalised treatment strategies for AD is highly important [2].

A rule-based treatment algorithm for AD management and control was found effective in a randomised controlled trial [3]. However, rule-based algorithms cannot recommend the treatment that is most likely to be effective for a given patient as they cannot simulate the outcomes of multiple regimens.

In this study, we aim to propose a computational pipeline that can generate personalised treatment recommendations for AD. We will achieve it by predicting how eczema severity on the following day will change if different treatment regimens are applied, e.g. if the patient applies topical corticosteroids and/or emollients today. The pipeline will recommend treatment regimens that will reduce the eczema severity predicted for the next day.

Our proposed pipeline is based on EczemaPred [4], a computational framework to model dynamic changes in eczema severity. EczemaPred consists of a collection of Bayesian state-space models that can predict the evolution of individual severity items (e.g. dryness, redness) constitutive of severity scores. Aggregating the predicted scores for relevant severity items provides us with the prediction of eczema severity scores. EczemaPred models address multiple challenges when working with real-world eczema severity data and can quantify uncertainty in severity measurement and prediction. The effectiveness of EczemaPred was previously demonstrated for predicting the evolution of Patient-Oriented SCORing Atopic Dermatitis (PO-SCORAD) [5]. Here we use EczemaPred models to estimate the efficacy of topical corticosteroids and emollient cream and produce treatment recommendations by applying Bayesian decision theory [6].

## METHODS

We developed a computational pipeline to generate personalised treatment recommendations (Fig. 1A). The central part of the pipeline is a multivariate Bayesian state-space generative model based on EczemaPred (Fig. 1B, detailed in the Model subsection below). The model’s inputs are PO-SCORAD assessed daily by patients, SCORAD evaluated monthly by clinical staff, and treatment usage (yes/no) within the past two days (grey circles in Fig. 1B). We also used knowledge from a prior study in the form of power priors to inform the parameters of the EczemaPred model to reach more robust conclusions and kickstart model training. The model produces inferences about treatment effects, the dynamics of the severity and its measurement biases, as well as predictions for future PO-SCORAD and SCORAD scores. The algorithm recommends whether or not to use topical corticosteroids, emollient creams, both or none on a given day to reduce predicted AD severity for the next day.

**Figure 1:**
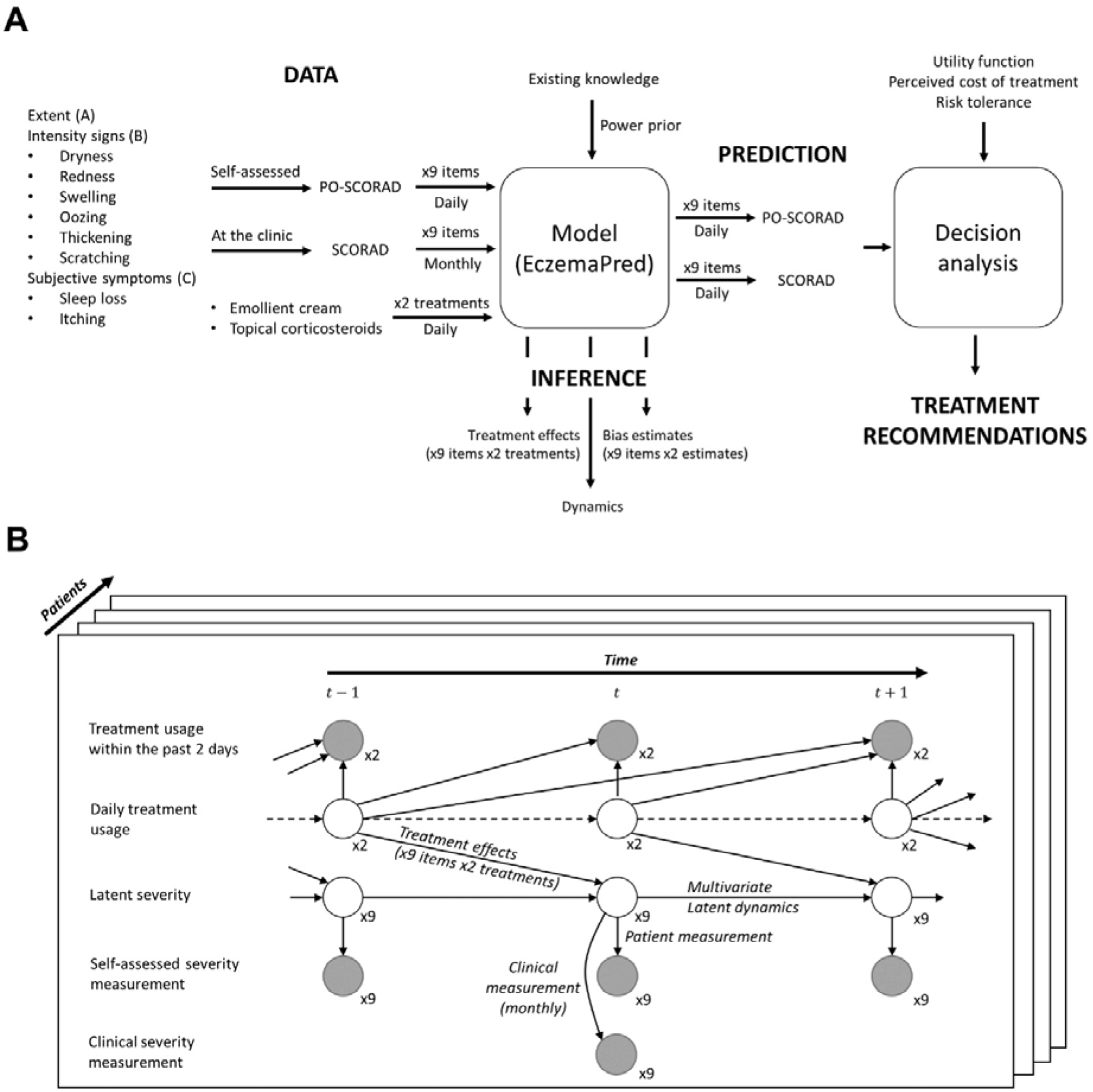
A) Overview of the method. The data (PO-SCORAD, SCORAD, treatment usage) and existing knowledge (power prior) are inputs to the model to produce inferences about treatment effects, the dynamics of the latent severity, and biases between SCORAD and PO-SCORAD. Treatment recommendations are generated from predictions on PO-SCORAD and SCORAD for a given utility function. B) Schematic of the EczemaPred model. Grey and white circles correspond to observed and latent variables, respectively

**Figure 1:**
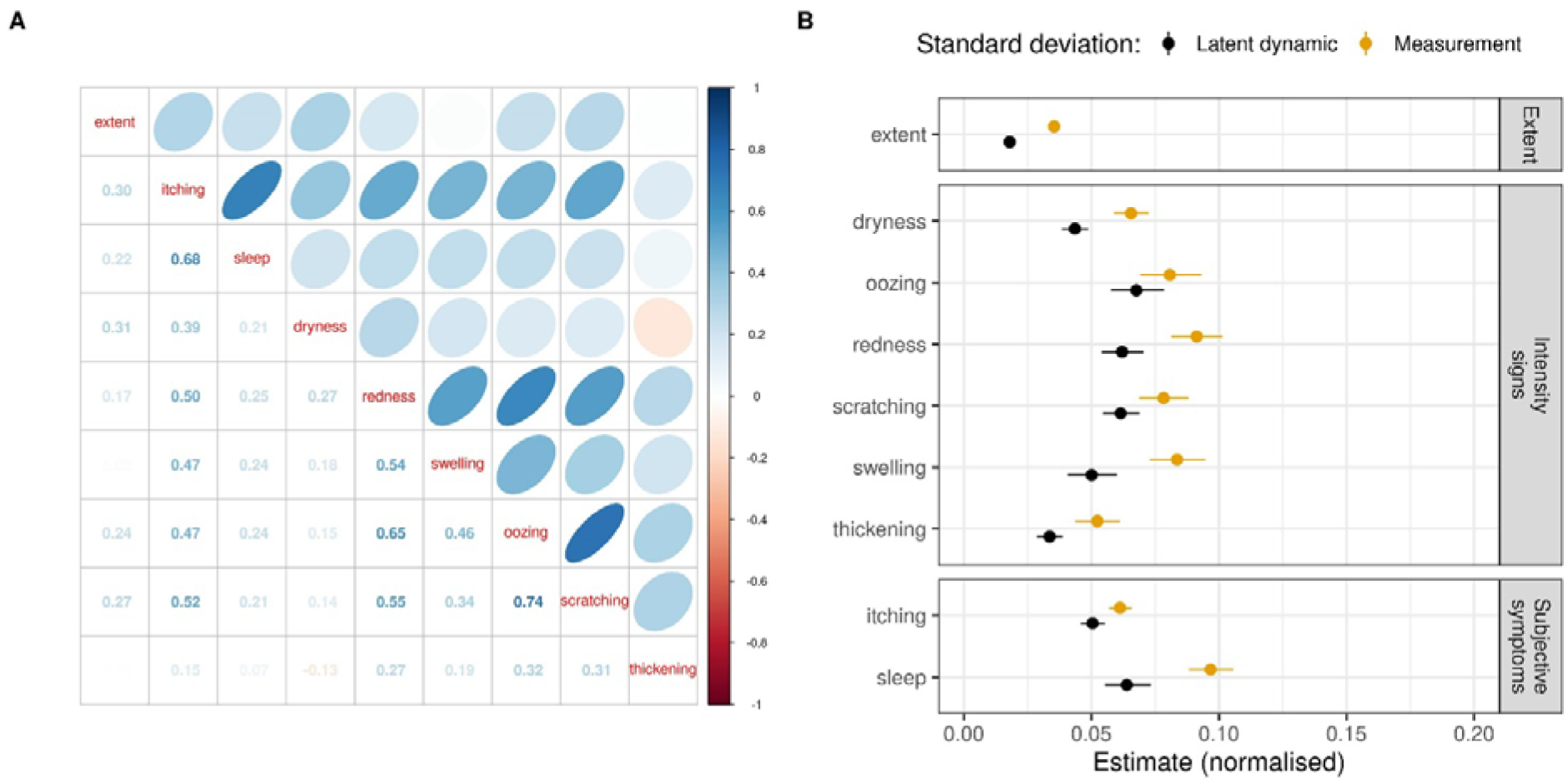
Posterior estimates of the state-space model. A) The expected correlation of the changes between the nine latent severity items. An ellipse in the upper diagonal matrix represents the strength of the correlations. The lower diagonal matrix displays the expected correlation coefficients. B) Estimates of the measurement and latent dynamic standard deviations for all severity items (normalised by the range of the score, mean and 90% credible intervals).

### Data

We used the previously published data from an observational study (ClinicalTrials.gov, NCT04553224) with 16 adult AD patients (mean age 25 years, SD=5) with a mean SCORAD of 34.6 (SD=4.4) at inclusion [4]. The study was approved by IEC (CPP Ile de France V, Saint Antoine Hospital, n°582211) and took place at Hôtel Dieu Hospital in Toulouse, France. The study participants gave written informed consent. Patients recorded PO-SCORAD daily using an app (https://www.poscorad.com) for up to 12 weeks (84 days), resulting in 1136 patient-day observations (13.6% missing values between the first and last observations). They also recorded whether treatment was used within the past two days in the daily app, for topical corticosteroids and emollient cream. During the study period, SCORAD was measured by a trained clinical staff every four weeks. Example data from a representative patient is shown in Fig. 2.

**Figure 2:**
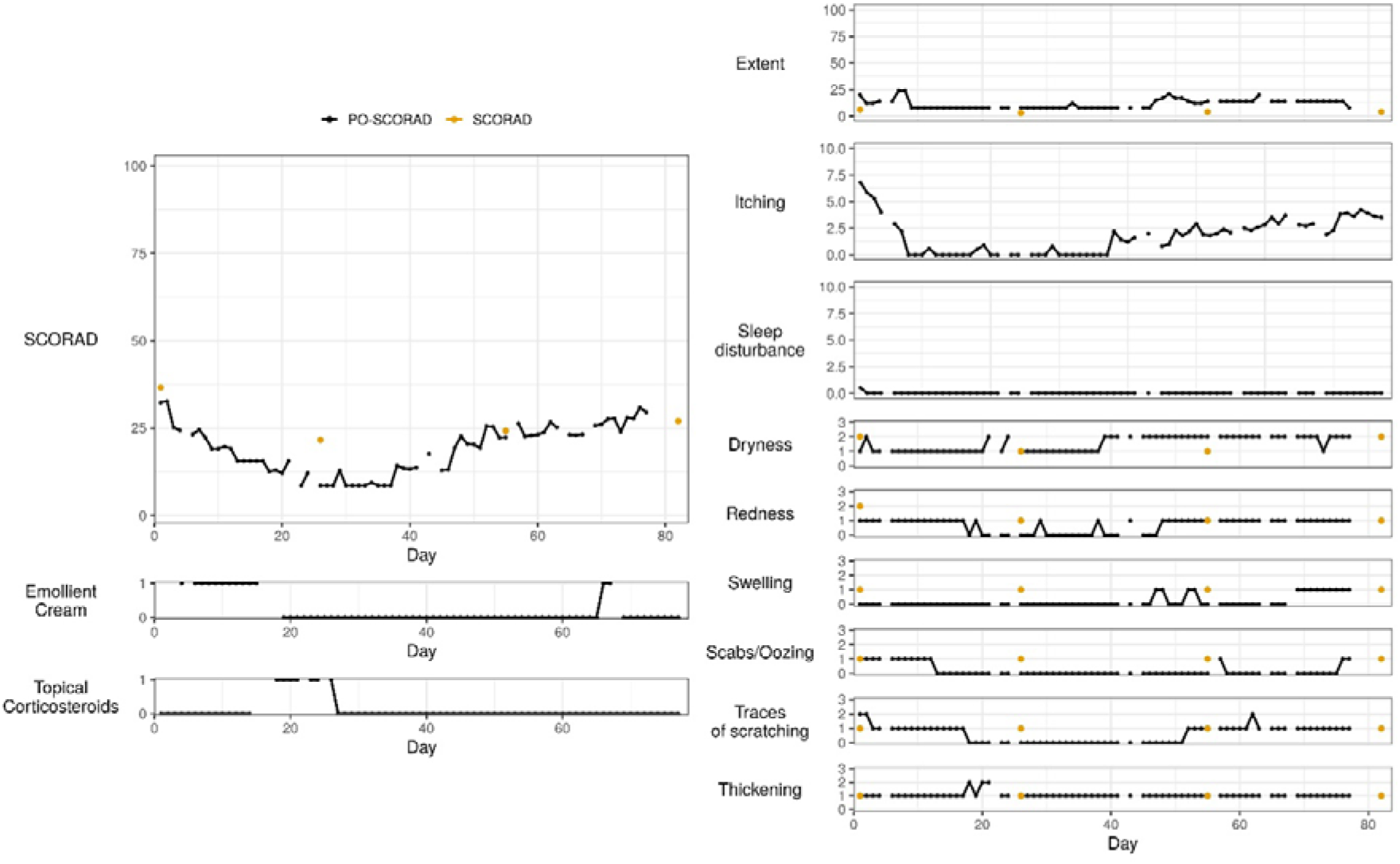
Data from a representative patient. Left: Trajectories of the aggregate PO-SCORAD severity score, the usage of emollient cream and topical corticosteroid within the past two days. Right: Trajectories of the nine severity items of PO-SCORAD. Orange dots correspond to the SCORAD measured every four weeks. The lines are broken when the measurements are missing.

**Figure 2:**
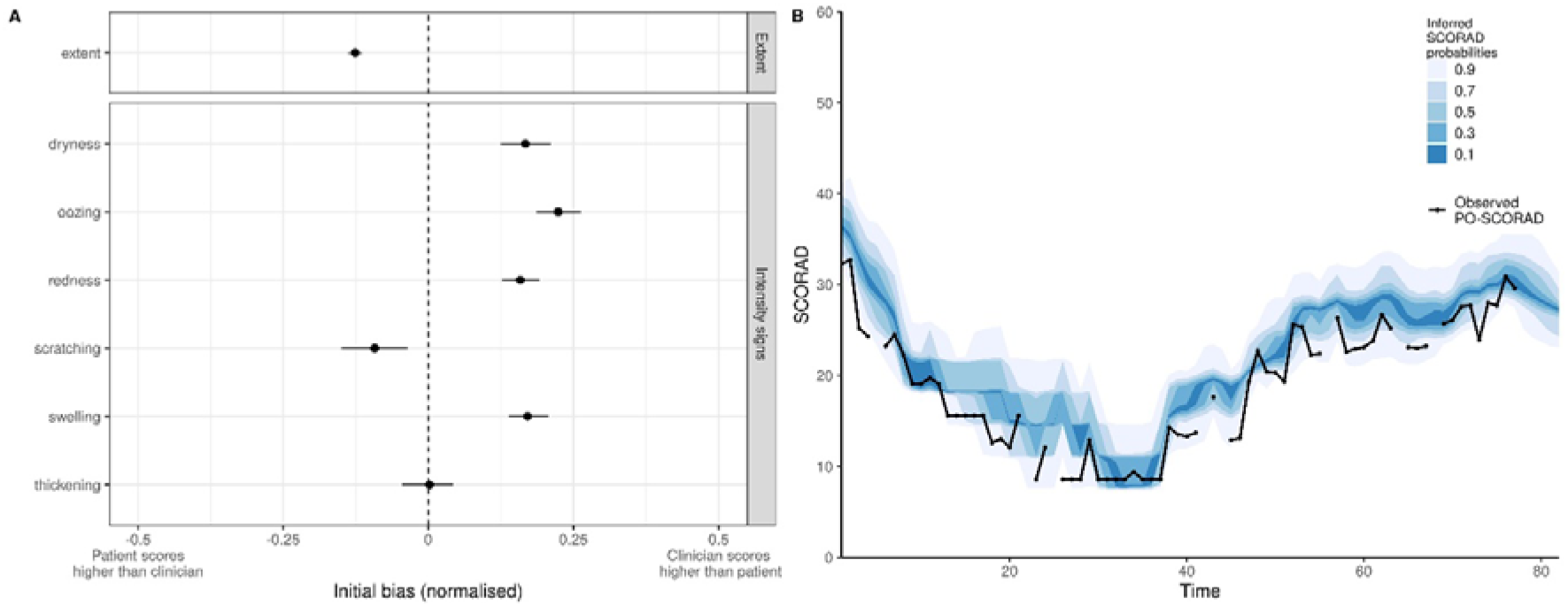
Calibration of PO-SCORAD measurements using SCORAD. A) Estimates of the initial bias (at day one) for SCORAD relative to PO-SCORAD (mean and 90% credible intervals; positive bias means SCORAD > PO-SCORAD), normalised by the range of the score. For example, patients are expected to overestimate the extent (with the range of [0, 1000]) by 0.13 × 100 =13 compared to the score given by clinicians. B) PO-SCORAD trajectories and the corresponding estimates of SCORAD a posteriori. The distribution is represented by stacked credible intervals in shades of blue. For this representative patient, estimated SCORAD (i.e. what a clinician would have measured) would be higher than PO-SCORAD (assessed by the patient), on average.

SCORAD (and PO-SCORAD by extension) is defined by *0*.*2 A + 3*.*5 B +C*, where *A* ∈ [0, 100] corresponds to the extent (the percentage of the area affected by eczema in the whole body, taking discrete values in the dataset), ∈ [0, 18] to intensity signs and *C* ∈ [0, 20] to subjective symptoms. The intensity signs component (*B*) is the sum of the scores for six intensity signs (dryness, redness, swelling, oozing, scratching, and thickening), each of which is assessed on an ordinal scale as 0 (absent), 1 (mild), 2 (moderate) or 3 (severe). The subjective symptoms component (*C*) is the sum of scores for two symptoms (itching and sleep loss), each of which is assessed on a visual analogue scale from 0 (no symptom) to 10 (severe symptom), taking discrete values with a resolution of 0.1 in the dataset (0.0, 0.1, …, 10.0). In this study, we work with the nine severity items of SCORAD (extent, six intensity signs and two subjective symptoms) rather than the aggregate score to extract as much information as possible from the data.

### Model

We used the previously published EczemaPred PO-SCORAD model [4] and extended it by integrating clinical measurements (SCORAD) and treatment usage data (Fig. 1B). The original EczemaPred PO-SCORAD model consisted of nine independent Bayesian state-space models, one for each severity item. The state-space models assume the observed severity items are the imperfect measurement of a latent severity (representing a “true severity”) which we assumed to follow a random walk.

In this paper, we modelled the severity items jointly by assuming multivariate latent dynamics, i.e. changes in the latent severity items are correlated. We modelled the measurement of all severity items using ordinal logistic distributions, which provide a way to control the variance/precision of the measurements. We proposed a parametrisation of the ordinal logistic distribution that is more interpretable and scales better to a higher number of categories, so that it can be used to model extent and subjective symptoms, instead of using Binomial distributions as in [4].

We integrated SCORAD measurements in the model by assuming that they are derived from the same latent score as for PO-SCORAD items but with a measurement bias between PO-SCORAD measured by patients and SCORAD measured by clinical staff. The bias is severity item-dependent and can decrease with time because PO-SCORAD assessments were found to become closer to SCORAD with experience [5]. We also assumed that SCORAD measurements are more precise (with a smaller variance for the measurement error) than PO-SCORAD measurements. We did not calibrate subjective symptoms as they are common for PO-SCORAD and SCORAD.

We included trend and treatment response components in the latent dynamics of each severity item. The trend component corresponds to an exponential smoothing of the difference between latent severity items at consecutive times. For the treatment response component, we first deconvolved the time-series data of treatment usage within the past two days, using deterministic and probabilistic inference, to obtain a time-series of daily treatment usage. The inferred daily treatment usage is then used to model item-dependent treatment effects for corticosteroids and emollients, assuming treatment usage at day only influences the severity at day *t* + 1.

Details of the model, including equations, are shown in Supplementary A.

### Priors

We used a power prior for the model parameters corresponding to the measurement and latent dynamics [7]. The power prior is an informative prior constructed from historical data. Informative priors are useful when working with small data and can help kickstart the model’s training as the model is “pre-trained”. To construct the power prior, we used the data from an already published study investigating the role of an emollient in 337 children with AD [8]. The data was also used to fit EczemaPred models [4]. The power prior was derived from the marginal posterior estimates of the population parameters of state-space models with ordinal logistic measurements and latent random walk.

Details of the power prior and priors for other parameters are given in Supplementary B.

### Treatment recommendation

Having developed a generative Bayesian model that can make severity predictions (for PO-SCORAD, SCORAD and their constitutive items) under different treatment conditions (actions), we applied Bayesian decision analysis to generate treatment recommendations under uncertainty that balance the costs and benefits of treatment use [6]. In Bayesian decision analysis, we choose a utility function that quantifies the “value” of taking a particular action (e.g. application of corticosteroids), make predictions corresponding to different actions, and recommend the action that maximises the expected utility (objective function) of the associated predictions. The objective function can include a risk-sensitive criterion to balance the benefit of the action (expected utility) and its risk (variance of utility, i.e. its uncertainty), which is also a way to balance the exploration-exploitation trade-off. We assumed a patient could be risk-averse (penalising uncertainty, or pessimistic), risk-neutral, or risk-seeking (welcoming uncertainty, or optimistic).

We used a simple utility function, *U* (*y* (*a*), *a*)= -(*y* (*a*) + *cost* (*a*)), where *a* denotes the action of using/not using topical corticosteroids or emollient creams, y(*a*) is the predicted SCORAD for the following day when taking action *a*, and *cost* (*a*) corresponds to the “perceived” cost of action *a*. The “perceived” cost could represent the fear of side effects [9], the inconvenience or monetary cost of using treatment, and, more generally, any mechanisms that drive poor adherence. For example, if the cost of not using treatment is 0 and the cost of using corticosteroids is 1, a risk-neutral patient would only use corticosteroids if the expected SCORAD after taking corticosteroids is 1 point less than the expected SCORAD when using no treatment.

We generated treatment recommendations by successively training the model every day, considering different “perceived” costs of using treatment (no cost, normal cost or high cost) and tolerance to risk (risk-averse, -neutral or -seeking). More details are given in Supplementary C.

### Model inference and validation

Model inference was performed using the Hamiltonian Monte Carlo algorithm in the probabilistic programming language Stan [10] with four chains and 2000 iterations per chain, including 50% warm-up. Prior predictive checks and fake data checks were conducted.

Our base model was the EczemaPred model with ordinal logistic measurement distributions for all severity items, as proposed in [4]. We evaluated the contribution of each new model component to the model predictive performance in a stepwise manner, by successively adding the power prior, the correlations between severity items, the calibration of PO-SCORAD with SCORAD, treatment effects and the dynamic trend of severity to the base model.

Comparison of the base model with standard time-series forecasting models was already conducted for all severity items and aggregate scores in [4] and was not repeated here, although we reported the performance of uniform and historical forecasts as references. Predictions were generated in a forward chaining setting where the model was retrained every four days and were evaluated with the logarithmic scoring rule (log predictive density, lpd) [11].

## RESULTS

We fitted the models to the data without finding any evidence for an absence of convergence by monitoring trace plots and checking the potential scale reduction factor 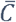 [12]. We conducted posterior predictive checks and found no clear discrepancies between the data and the models’ simulations.

### Model inference

#### (1) Dynamics of severity items

Joint fitting of the nine latent severity items of PO-SCORAD to our EczemaPred model revealed that changes in the severity items were positively correlated (Fig. 3A). This implies more uncertainty in the prediction of PO-SCORAD compared to when the severity items are independent, as changes accumulate rather than cancel out when we aggregate the severity items to derive PO-SCORAD. In particular, changes in scratching, oozing, and redness appear to be strongly correlated, and changes in itching are moderately correlated with changes in all intensity signs. Extent is only mildly correlated with subjective symptoms or intensity signs.

**Figure 3:**
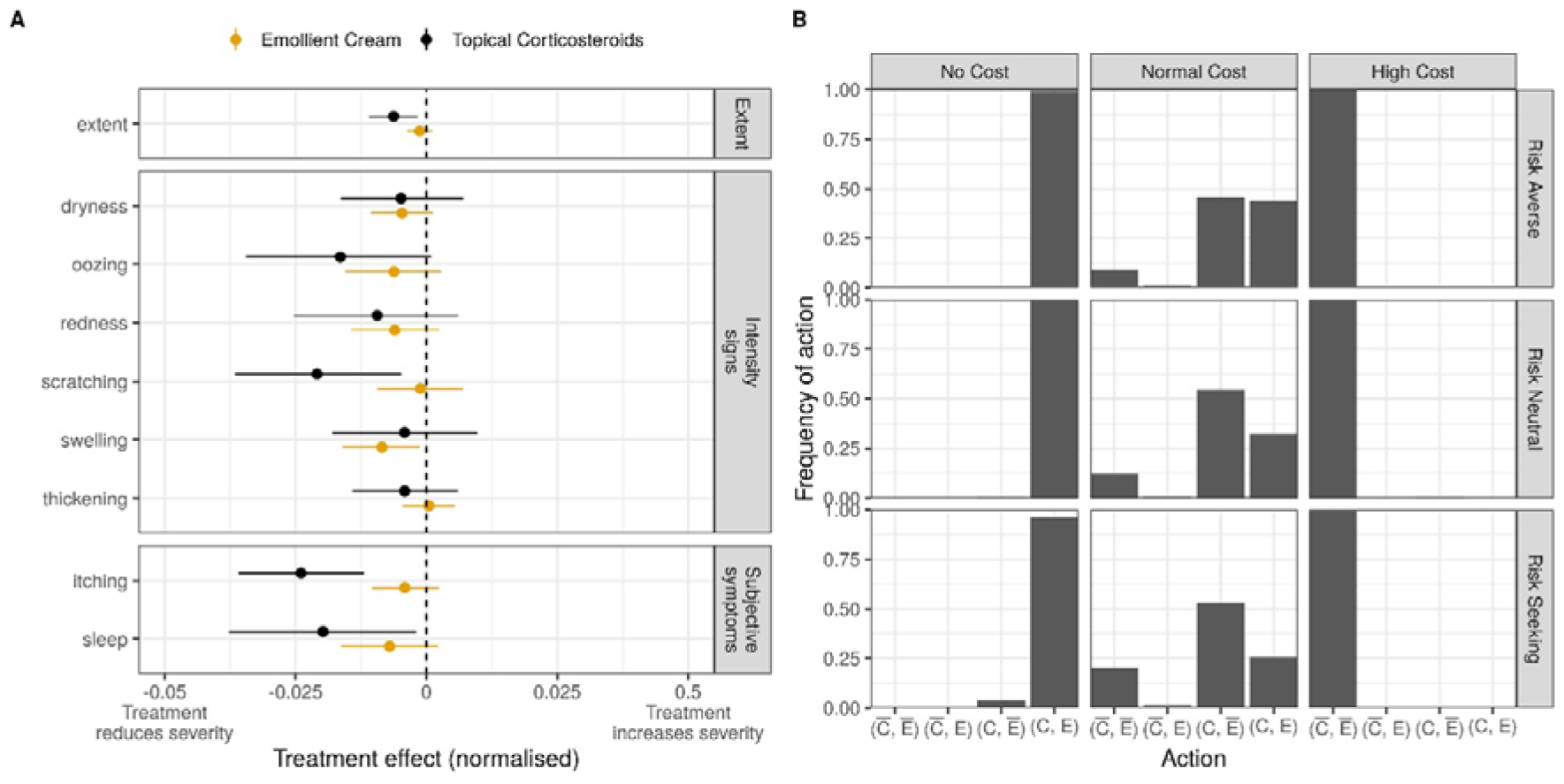
A) Average treatment effects on each severity item for topical corticosteroids (black) and emollient cream (orange), normalised by the range of the score (mean and 90% CI). For example, the expected effect of corticosteroids on the latent sleep loss item, defined in [0, 10], is -0.02, meaning that taking corticosteroids will decrease the sleep loss score by 0.02 × 10 = 0.20 on average. B) Distribution of recommended actions for no/normal/high perceived cost of treatment (vertical facets) and a risk-averse/neutral/seeking patient (horizontal facets). C and 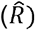 correspond to the action of using and not using corticosteroids, respectively; E and Ē correspond to the action of using and not using emollients, respectively.

The evolution of some items is more uncertain than others (Fig. 3B). For example, the evolution of oozing is more uncertain than the evolution of thickening. Most of the prediction uncertainty can be explained by the uncertainty of the measurement process, as the uncertainty of the measurement process is always larger than that of the latent dynamics (Fig. 3B). This highlights the difficulty in accurately modelling the evolution of AD severity.

#### (2) Calibration of PO-SCORAD with SCORAD

We estimated the measurement bias between patient-assessed PO-SCORAD and clinician-assessed SCORAD in our model. The direction and amplitude of the biases were item-dependent (Fig. 4A). For example, patients tend to overestimate the extent and scratching, but underestimate dryness, redness, swelling, and oozing, compared to clinicians, on average. The biases mostly stayed constant over time, except for scratching, for which the bias is nearly 0 after the second measurement at week 4 (Fig. S3). We would have expected the biases to decrease with time if patients were getting better at assessing their symptoms. It is possible that the patients in this dataset may have already been familiar with PO-SCORAD assessments or that learning did not happen without any feedback provided.

Using the estimated measurement biases, we converted PO-SCORAD predictions into SCORAD predictions (i.e. forecasting SCORAD) and inferred SCORAD values as if they had been measured daily (i.e. backcasting and nowcasting SCORAD, Fig. 4B). The severity trajectory for a representative patient (Fig. 4B) demonstrates that the expected SCORAD would have been higher than the observed PO-SCORAD. This is consistent with our estimates that clinicians tend to score intensity signs higher than patients (Fig. 4A) and with the fact that the intensity signs component (3.5*B*) contributes more to the total SCORAD than the extent component (0.2*A*).

#### (3) Treatment effects

The model parameters corresponding to treatment effects were estimated as negative, confirming that the application of treatment is associated with a decrease in AD severity (Fig. 5A). Topical corticosteroids were more effective than emollient creams in reducing AD severity on the following day, although estimates of treatment effects were small in absolute values. Treatment effects were also uncertain with a large CI, probably because nearly half of the patients (7/16, when excluding missing values in treatment usage) reported never using corticosteroids or emollients. In addition, treatment effects were highly heterogeneous across severity items. For example, a patient with severe scratching but no thickening of the skin would respond more to corticosteroids than a patient with no scratching but severe thickening, all else being equal.

### Treatment recommendations

We generated treatment recommendations for different decision profiles defined by no/normal/high perceived cost of treatment and a risk-averse/neutral/seeking nature of the patient (Fig. 5B). We confirmed a high perceived cost of treatment is associated with no treatment being recommended. In contrast, a null perceived cost of treatment is associated with both treatments being recommended, which was anticipated considering that application of either treatment is associated with a decrease in AD severity (Fig. 5A). The most recommended action for a “normal” perceived cost of treatment is to use corticosteroids but not emollients. This is consistent with the result that corticosteroids are more effective than emollients (Fig. 5A), as the benefit of additional use of emollients may not be worth their “perceived” cost in the “normal” cost scenario.

For a “normal” perceived cost of treatment, the algorithm is more likely to recommend “using both treatments” for a risk-averse patient (who penalises uncertainty in the outcome) than for a risk-seeking patient (who welcomes uncertainty in the outcome), for whom the algorithm recommends “using no treatments” more often because taking treatment does not sufficiently improve the best-case scenario for the cost of taking the treatment.

It is worth emphasising that recommendations can be “personalised” even though treatment parameters are not patient-dependent. Different actions can be recommended for patients with the same SCORAD (Fig. S4B) because treatment responses depend on the composition of severity items (Fig. 5A) i.e. the clinical phenotype. For example, application of treatments (corticosteroids and/or emollients) is recommended more often than no application when the severity is high (Fig. S4B), even though this is not explicitly implemented in the utility function. This may be a side effect of more severity items being present for severe AD and each severity item triggering treatment, resulting in more potential for improvement and overall better responses to treatment. The severity-dependence of the recommendations could also be confounded by the fact that patients tend to have a higher severity at the beginning of the study when the algorithm recommends “using both treatments” more (Fig. S4A). This also illustrates how recommendations can change as more data comes in, and treatment effects are learnt. These possible explanations highlight the difficulty of interpreting the descriptive summaries of recommendations post-hoc. Nonetheless, every recommendation can be transparently explained by examining the utility function of the patient and their tolerance to risk.

### Model validation

We validated the model to assess whether its new features (compared to the previously published EczemaPred models) were associated with improvements in predictive performance (Fig. S5). We did not find evidence that using additional information (power priors, SCORAD measurements, treatments) was associated with noticeable long-term improvements in predictive performance. This may not be surprising as the main contribution of the power prior is to accelerate the learning process, the addition of at most four SCORAD measurements is unlikely to change the performance of 12-week-long daily time-series by much, and treatment effects were expected to be small [13]. Similarly, modelling the trend in the latent dynamics did not improve performance, as the dynamic trend was estimated to be null (Fig. S2). More surprisingly, modelling correlations between severity items did not significantly change the average predictive performance measured by the lpd, although the resulting predictive distributions were different.

#### DISCUSSION

In this study, we developed a computational pipeline to generate personalised treatment recommendations for AD (Fig. 1). The pipeline integrates multi-dimensional data (Fig. 2) and uses EczemaPred [4] to predict the evolution of eczema severity and infer treatment effects. Our model demonstrated that changes in eczema severity items are positively correlated (Fig. 3A). We estimated the effects of using topical corticosteroids and emollient creams on AD severity and demonstrated a proof-of-concept to generate personalised recommendations (Fig. 5). To support treatment decision, we calibrated self-assessed PO-SCORAD using SCORAD assessed by clinical staff (Fig. 4). We also leveraged existing knowledge about the dynamics of the disease by designing an informative prior derived from historical data to accelerate model training (Fig. S1).

This study clarified the strong heterogeneity in the latent dynamics of severity, in the biases between self-assessed and clinician-assessed severity, and in treatment effects. For example, we found that patients are expected to overestimate the extent but underestimate redness, and that topical steroids are more effective in reducing scratching than skin thickening (lichenification). As a result, treatment effects measured with an aggregate severity score (e.g. SCORAD) may appear different between two patients with the same score but with distinct clinical phenotypes (e.g. measured in PO-SCORAD with different values of extent or intensity of signs like scratching or thickening), even though treatment effects are not patient-dependent per se [14]. The fact that the clinical phenotype of patients may confound treatment effects highlights the importance of modelling severity items rather than the aggregate scores.

We showed how EczemaPred can be used to simultaneously make predictions, inferences about treatment effects, and treatment recommendations. Using the same model ensures producing consistent results when addressing several research questions. For example, our treatment recommendations are consistent with the inferred treatment effects and the model’s predictions for PO-SCORAD and SCORAD. This would not necessarily be the case if we used different models/analytical methods for score predictions and treatment recommendations, or interpreted the decisions of a black-box treatment recommendation algorithm a posteriori where model explanations may not reflect their actual decision process [15]. The recommendations also account for the correlated dynamics between severity items, and benefit from the knowledge of prior studies, while considering uncertainty in measurements, parameters and predictions.

For medical decision-making, we usually have only imperfect data but the decisions cannot be postponed until more evidence is collected. Under such circumstances, it is crucial to have a flexible model that can integrate all the available information [16]. For example, PO-SCORAD (assessed by patients) is supposedly less accurate than SCORAD (assessed by trained clinical staff) but can be measured daily, unlike SCORAD which can only be measured infrequently when patients visit a clinic. By calibrating PO-SCORAD with SCORAD, we can get the best of high-quality (SCORAD) and high-frequency (PO-SCORAD) measurements to improve the training data quality and build trust in the model’s output. Using informative priors from historical data also mitigates the cold-start problem.

This study has some limitations. Our proposal to generate treatment recommendations is very much a proof-of-concept. Even if the recommendations were optimal and reliable, the recommended treatments would only be associated with small improvements in AD severity because the estimated treatment effects are small and uncertain. More importantly, the model is not causal. This is why we did not attempt to evaluate the quality of the recommendations. Potentially many confounding factors are missing, making counterfactual inference not possible. For example, better quality treatment data would be required, such as the daily usage, potency and quantity of treatment applied. It is also possible that the self-reported treatment usage may sometimes be inaccurate. The suggested recommendations are illustrative, as the utility function and decision parameters would need to be adjusted to match patients’ preferences and could evolve over time. We did not detect any improvement in the predictive performance despite the additional complexity implemented in the model, highlighting the difficulty of accurately predicting the evolution of eczema severity. Data from a larger cohort would be required to investigate patient-dependence (in treatment effects, measurement biases, or even dynamical parameters) and to ensure our results can be generalised. It would also be relevant to include additional treatments such as topical calcineurin inhibitors and PDE4 inhibitors in the data.

In a clinical setting, our model could be used to help track the evolution of AD severity scores between two visits with great accuracy, by removing the biases of self-assessed severity measures. The model could also provide insights into the evolution of the disease and be a tool to initiate a discussion between clinicians and patients. The decision to initiate treatment could be fully data-driven with prior knowledge (e.g. from similar patients) being incorporated systematically. Switching from non-effective treatment could also be sped up compared to standard care because the model would continuously refine its estimate of treatment effects. Such an algorithm would therefore work as if patients were in their own adaptive clinical trial, and personalised treatment recommendation algorithms could improve the care of AD patients in future.

As a future direction, the proposed model could be used to reduce the dimensionality of the latent severity space to search for common patterns in the severity trajectories and cluster patients into different endotypes [17]. We can hypothesise that the correlated latent trajectories associated with different severity items may be partly redundant and the manifestation of a few potentially independent mechanisms that could stratify patients. This type of model-based clustering has already been applied to investigate whether the “atopic march” hypothesis was supported by data [18]. Data-driven modelling may be more useful if applied to better understand AD dynamics than for prediction only, considering the difficulty in accurately predicting AD severity’s evolution.

## Data Availability

The analysis code is available at https://github.ic.ac.uk/tanaka-group/EczemaTreat

## Funding acknowledgement

This study was funded by the British Skin Foundation (005R18). Funders had no involvement in study design.

## REFERENCES

[1] S. M. Langan, A. D. Irvine, and S. Weidinger, “Atopic dermatitis” The Lancet, vol. 396, no. 10247, pp. 345–360, Aug. 2020, 10.1016/S0140-6736(20)31286-1.

[2] S. J. Galli, “Toward precision medicine and health: Opportunities and challenges in allergic diseases,” Journal of Allergy and Clinical Immunology, vol. 137, no. 5, pp. 1289–1300, May 2016, 10.1016/j.jaci.2016.03.006.

[3] J. Schmitt, M. Meurer, U. Schwanebeck, X. Grählert, and K. Schäkel, “Treatment following an evidence-based algorithm versus individualised symptom-oriented treatment for atopic eczema: A randomised controlled trial,” Dermatology, vol. 217, no. 4, pp. 299–308, 2008, 10.1159/000151355.

[4] G. Hurault et al., “EczemaPred: A computational framework for personalised prediction of eczema severity dynamics,” Clinical and Translational Allergy, vol. 12, no. 3, pp. e12140, Mar. 2022, 10.1002/clt2.12140.

[5] J. F. Stalder et al., “Patient-Oriented SCORAD (PO-SCORAD): A new self-assessment scale in atopic dermatitis validated in Europe,” Allergy: European Journal of Allergy and Clinical Immunology, vol. 66, no. 8, pp. 1114–1121, Aug. 2011, 10.1111/j.1398-9995.2011.02577.x.

[6] J. O. Berger, Statistical Decision Theory and Bayesian Analysis. New York, NY: Springer New York, 1985. 10.1007/978-1-4757-4286-2.

[7] J. G. Ibrahim, M. H. Chen, Y. Gwon, and F. Chen, “The power prior: Theory and applications,” Statistics in Medicine, vol. 34, no. 28, pp. 3724–3749, 2015, 10.1002/sim.6728.

[8] G. S. Tiplica et al., “The regular use of an emollient improves symptoms of atopic dermatitis in children: a randomized controlled study,” Journal of the European Academy of Dermatology and Venereology, vol. 32, no. 7, pp. 1180–1187, Jul. 2018, 10.1111/jdv.14849.

[9] A. W. Li, E. S. Yin, and R. J. Antaya, “Topical Corticosteroid Phobia in Atopic Dermatitis,” JAMA Dermatology, vol. 153, no. 10, p. 1036, Oct. 2017, 10.1001/jamadermatol.2017.2437.

[10] B. Carpenter et al., “Stan□: A Probabilistic Programming Language,” Journal of Statistical Software, vol. 76, no. 1, pp. 1–32, 2017, 10.18637/jss.v076.i01.

[11] A. Gelman, J. Hwang, and A. Vehtari, “Understanding predictive information criteria for Bayesian models,” Statistics and Computing, vol. 24, no. 6, pp. 997–1016, Nov. 2014, 10.1007/s11222-013-9416-2.

[12] A. Vehtari, A. Gelman, D. Simpson, B. Carpenter, and P.-C. Bürkner, “Rank-Normalization, Folding, and Localization: An Improved R for Assessing Convergence of MCMC (with Discussion),” Bayesian Analysis, vol. 16, no. 2, Jun. 2021, 10.1214/20-BA1221.

[13] G. Hurault, E. Domínguez-Hüttinger, S. M. Langan, H. C. Williams, and R. J. Tanaka, “Personalized prediction of daily eczema severity scores using a mechanistic machine learning model,” Clinical and Experimental Allergy, vol. 50, no. 11, pp. 1258–1266, Aug. 2020, 10.1111/cea.13717.

[14] S. Senn, “Statistical pitfalls of personalized medicine,” Nature, vol. 563, no. 7733, pp. 619–621, Nov. 2018, 10.1038/d41586-018-07535-2.

[15] C. Rudin, “Stop explaining black box machine learning models for high stakes decisions and use interpretable models instead,” Nature Machine Intelligence, vol. 1, no. 5, pp. 206–215, May 2019, 10.1038/s42256-019-0048-x.

[16] A. E. Ades and A. J. Sutton, “Multiparameter evidence synthesis in epidemiology and medical decision-making: current approaches,” Journal of the Royal Statistical Society: Series A (Statistics in Society), vol. 169, no. 1, pp. 5–35, Jan. 2006, 10.1111/j.1467-985X.2005.00377.x.

[17] T. Bieber et al., “Clinical phenotypes and endophenotypes of atopic dermatitis: Where are we, and where should we go?,” Journal of Allergy and Clinical Immunology, vol. 139, no. 4, pp. S58–S64, Apr. 2017, 10.1016/j.jaci.2017.01.008.

[18] D. C. M. Belgrave et al., “Developmental Profiles of Eczema, Wheeze, and Rhinitis: Two Population-Based Birth Cohort Studies,” PLoS Medicine, vol. 11, no. 10, p. e1001748, Oct. 2014, 10.1371/journal.pmed.1001748.

