## Supplementary material for "Data-driven personalised recommendations for eczema treatment using a Bayesian model of severity dynamics"

**Supplementary Materials**

#### Notation

We will use the following conventions:

- Vectors are marked in bold.
- Matrices are denoted with a capital letter.
- Subscripts are used to subset vectors or matrices.
- Superscripts in parenthesis are used to index patients.
- $*$ denotes the element-wise multiplication.
- $y \sim \mathcal{N}(\mu, \sigma^2)$ means that $y$ is normally distributed with mean $\mu$ and variance $\sigma^2$. Similarly, $\boldsymbol{y} \sim \mathcal{N}(\boldsymbol{\mu}, \Sigma)$ means that the vector $\boldsymbol{y}$ follows a multivariate normal distribution with mean $\boldsymbol{\mu}$ and covariance $\Sigma$.

We will use the following notation:

- $\boldsymbol{y}^{(k)}(t)$ is the vector of measured PO-SCORAD items for the $k$-th patient at time $t$ (dimension $9 \times 1$).
- $\boldsymbol{q}^{(k)}(t)$ is the vector of measured SCORAD items for the $k$-th patient at time $t$ (dimension $9 \times 1$).
- $\hat{\boldsymbol{y}}^{(k)}(t)$ is the vector containing the latent score for each severity item for the $k$-th patient at time $t$ (dimension $9 \times 1$).
- $\hat{\boldsymbol{u}}^{(k)}(t)$ is the vector containing the probability that treatment was used (0 or 1 when inferred deterministically, otherwise unknown), for each treatment, for the $k$-th patient at time $t$ (dimension $2 \times 1$).
- $\boldsymbol{b}^{(k)}(t)$ is the vector containing the trend for each latent score for the $k$-th patient at time $t$ (dimension $9 \times 1$).
- $\boldsymbol{M}$ such that the $i$-th severity item takes values in $[0, M_i]$ where $M_i = 3$ for intensity signs and $M_i = 100$ for extent and subjective symptoms (subjective symptoms have a resolution of 0.1 so are scaled to take integer values in $[0, 100]$ and scaled back to $[0, 10]$ for predictions).

### A. Model

We developed a Bayesian state-space model, which is an extension of the "OrderedRW" model introduced in [1].

#### A.1. Ordered logistic distribution

##### A.1.1. Previous parametrisation

In [1], we parametrised the ordered logistic distribution for a discrete ordinal outcome $y \in \{0, ..., M\}$ ($M + 1$ categories) by a location $\eta$ and an ordered vector of cut-off values/cutpoints $\boldsymbol{c} = (c_1 ... c_M)$ of size $M$ ($c_1 < c_2 < ... < c_M$), such as:

$$\text{OrderedLogistic}\left(y|\eta, \boldsymbol{c}\right) = \begin{cases} 1 - \text{logit}^{-1}\left(\eta - c_1\right) & \text{if } y = 0 \\ \text{logit}^{-1}\left(\eta - c_y\right) - \text{logit}^{-1}\left(\eta - c_{y+1}\right) & \text{if } 0 < y < M \\ \text{logit}^{-1}\left(\eta - c_M\right) & \text{if } y = M \end{cases} \quad (1)$$

The ordered logistic distribution can be understood as a logistic distribution (the distribution for which the cumulative distribution is a logistic function) with location $\eta$ and scale 1, being discretised using the cut-offs $\boldsymbol{c}$. As such, the probability of observing $y$ is equal to the area under the logistic distribution between the $y$-th and the $y+1$-th cut-offs[1].

In the parametrisation used in [1], the cutpoints are unknown, which implies that the range of the latent score ($c_M - c_1 = c_M$) varies depending on the measurement error of the severity item. As a result, it was difficult to interpret the latent scores, compare and quantify the amount of measurement error for different severity items. The distribution was also parametrised by the difference $\boldsymbol{\delta}$ between two consecutive cutpoints, which implies that the variance of the marginal distribution of cutpoints increased with the cutpoint index, even though there is no reason to expect this a priori.

##### A.1.2. New parametrisation

We propose to parametrise the ordered logistic distribution using a logistic distribution with an unknown standard deviation $\sigma$ (which is related to the scale $s$ of the logistic distribution by $\sigma = \frac{\pi}{\sqrt{3}} s$) and unknown cutpoints, but for which the range (difference between the first and last cutpoints) is fixed. The variance of the ordered logistic distribution in (1) is indeed controlled by the range of $\boldsymbol{c}$, as scaling $\eta$ and $\boldsymbol{c}$ by a scalar $\frac{1}{s}$, i.e. $\text{OrderedLogistic}(y|\frac{\eta}{s}, \frac{1}{s}\boldsymbol{c})$, is equivalent to thresholding a logistic

[1]The cut-offs vector $\boldsymbol{c}$ can be appended with $c_0 = -\infty$ and $c_{M+1} = +\infty$ for the sake of the argument.

distribution with location $\eta$ and scale $s$ by the vector $\boldsymbol{c}$.

For $y \in \{0, ...M\}$ ($M + 1$ categories):

$$\text{OrderedLogistic}\left(y|\eta, \sigma, \boldsymbol{\delta}\right) = \begin{cases} 1 - \text{logit}^{-1}\left(\frac{\eta - c_1}{s}\right), \text{if } y = 0, \\ \text{logit}^{-1}\left(\frac{\eta - c_y}{s}\right) - \text{logit}^{-1}\left(\frac{\eta - c_{y+1}}{s}\right), \text{for } 0 < y < M, \\ \text{logit}^{-1}\left(\frac{\eta - c_M}{s}\right), \text{if } y = M. \end{cases} \tag{2}$$

Where:

- $\boldsymbol{\delta}$ a simplex vector of size $M - 1$
- $\boldsymbol{c}$ a vector of size $M$ such as $c_1 = 0.5$ and $c_{i+1} = c_i + (M - 1)\delta_i$ for $i > 0$. Thus, $c_M = M - 0.5$.
- $\sigma$ is the standard deviation of the logistic distribution and $s = \sigma\frac{\sqrt{3}}{\pi}$ is the scale of the logistic distribution

Defining $\boldsymbol{c}$ as above implies that the range of $\eta$ is approximately the same as the range $[0, M]$ of the observations. Moreover, if cutpoints are equally spaced, the expected value of the distribution when $\eta \in \{1, ..., M - 1\}$ is equal to $\eta$.

We assumed a symmetric Dirichlet prior on $\boldsymbol{\delta}$, which specifies a joint prior on $\boldsymbol{c}$ to regularise the cutpoints, making the model scale well to a high number of categories:

$$\boldsymbol{\delta} \sim \text{Dirichlet}(\mathbf{2}) \tag{3}$$

And we assumed a lognormal prior on $\sigma$, which translates to a 95% CI that is approximately $[0.02M, 0.40M]$, thus allowing very precise or very imprecise measurement that covers the entire range of the score:

$$\frac{\sigma}{M} \sim \log \mathcal{N}\Big( - \log(10), \log(2)^2\Big) \tag{4}$$

#### A.2. Base model: ScoradPred

We define the base EczemaPred model for PO-SCORAD, that consists of independent state-space models with ordered logistic measurement distributions and random walk latent dynamics:

$$\begin{aligned} y_i^{(k)}(t) &\sim \text{OrderedLogistic}\left(\hat{y}_i^{(k)}(t), (\sigma_y)_i, \boldsymbol{\delta}_i\right) && (5) \\ \hat{y}_i^{(k)}(t+1) &\sim \mathcal{N}\big(\hat{y}_i^{(k)}(t), (\sigma_1)_i^2\big) && (6) \\ \hat{y}_i^{(k)}(t_0) &\sim \mathcal{N}\big((\mu_0)_i, (\sigma_0)_i^2\big) && (7) \end{aligned}$$

Where:

- $\boldsymbol{\sigma}_{\mathrm{y}}$ is the vector containing the standard deviation of the ordered logistic distributions (size $9$).
- $\boldsymbol{\delta}_i$ is the vector of cut-offs of the ordered logistic distribution for the $i$-th severity item (size $M_i - 1$).
- $\boldsymbol{\sigma}_{\mathrm{l}}$ is the vector containing the standard deviations of the latent dynamics (size $9$).
- $\boldsymbol{\mu}_{\mathbf{0}}$ is the vector of population means of the initial latent scores for each severity item (size $91$).
- $\boldsymbol{\sigma}_{\mathbf{0}}$ is the vector of population standard deviations of the initial latent scores for each severity item (size $9$).

#### A.3. Proposed model

We extend the base model by:

- Modelling correlations between changes in latent scores.
- Integrating clinical measurements (SCORAD) in addition to patient-assessed measurements (PO-SCORAD).
- Including treatment usage data to estimate treatment effects.
- Modelling the trend of latent severity items.

##### A.3.1. Latent dynamics

We assume that the evolution of $\hat{\boldsymbol{y}}$ follows a multivariate normal distribution, defining a vector autoregressive model:

$$\hat{\boldsymbol{y}}^{(k)}(t) \sim \mathcal{N}\big(\hat{\boldsymbol{y}}^{(k)}(t-1) + \boldsymbol{b}^{(k)}(t-1) + \Theta\hat{\boldsymbol{u}}^{(k)}(t-1), \Sigma\big) \tag{8}$$

Where:

- $\Sigma = \mathrm{diag}(\boldsymbol{\sigma}_{\mathrm{l}})\,\Omega\,\mathrm{diag}(\boldsymbol{\sigma}_{\mathrm{l}})$ is the $9 \times 9$ covariance matrix of the multivariate normal distribution.
- $\boldsymbol{\sigma}_{\mathrm{l}}$ is the vector of marginal standard deviations (for each severity item) of the multivariate normal distribution (size $9$).
- $\Omega$ is the $9 \times 9$ correlation matrix of the multivariate normal distribution.
- $\Theta$ is the $9 \times 2$ matrix containing the treatment effects of each treatment (columns) for each severity item (rows).

###### A.3.2. Initial conditions

We also assume that the initial conditions (at $t = t_0$) follow a multivariate normal distribution:

$$\hat{\boldsymbol{y}}^{(k)}(t_0) \sim \mathcal{N}\big(\boldsymbol{\mu_0}, \Sigma_0\big) \tag{9}$$

Where:

- $\boldsymbol{\mu_0}$ is the vector of population means of the initial latent scores for each severity item (size $9$).
- $\Sigma_0 = \operatorname{diag}(\boldsymbol{\sigma_0})\Omega_0 \operatorname{diag}(\boldsymbol{\sigma_0})$ is the $9 \times 9$ covariance matrix of the initial latent scores.
- $\boldsymbol{\sigma_0}$ is the vector of marginal population standard deviations of the latent scores for each severity item (dimension $9$).
- $\Omega_0$ is the $9 \times 9$ correlation matrix for the initial latent scores.

###### A.3.3. Trend

We model the trend of the latent scores using exponential smoothing [2]:

$$\boldsymbol{b}^{(k)}(0) = \mathbf{0} \tag{10}$$

$$\boldsymbol{b}^{(k)}(t) = \boldsymbol{\phi} * \big(\hat{\boldsymbol{y}}^{(k)}(t) - \hat{\boldsymbol{y}}^{(k)}(t-1)\big) + (\mathbf{1} - \boldsymbol{\phi}) * \boldsymbol{b}^{(k)}(t-1) \tag{11}$$

Where:

- $\boldsymbol{\phi}$ is the vector containing the smoothing trend parameter for each severity item (size $9$). If $\phi_i = 0$, then the trend is constant and equal to 0. If $\phi_i = 1$, then the trend is not smoothed.

We do not define a double exponential smoothing, as the measurement process already acts as if the latent scores were a smoothed version of the observations. Moreover, to keep things simple, we do not assume any damping or that the smoothing parameter is patient-dependent.

###### A.3.4. Daily treatment usage inference

The dataset contains the information on whether treatment “was used within the past two days", which we deconvolve to obtain daily treatment usage information.

For any of the two treatments (we drop the subscript $j$ indexing treatment out of concision), let:

- $v$ be the time-series of “treatment usage within the past two days".

- $u$ the time-series of daily treatment usage.
- then $\hat{u}(t) = P\big(u(t) = 1\big)$ the probability that treatment was used at time $t$.

We can use logic to deterministically infer some values of $\hat{u}$ given the time-series $v$. More specifically, we identify three cases where deterministic inference is possible:

$$v(t) = 0 \qquad\qquad\qquad\qquad \Rightarrow \quad \hat{u}(t-2) = \hat{u}(t-1) = \hat{u}(t) = 0 \tag{12}$$

$$v(t) = 0 \quad \& \quad v(t+1) = 1 \quad \Rightarrow \quad \hat{u}(t+1) = 1 \tag{13}$$

$$v(t) = 1 \quad \& \quad v(t+1) = 0 \quad \Rightarrow \quad \hat{u}(t-2) = 1 \tag{14}$$

The values of $\hat{u}(t)$ that cannot be inferred deterministically are treated as parameters to be inferred by the model, given the likelihood:

$$P\big(v(t) = 1\big) = 1 - \big(1 - \hat{u}(t)\big)\big(1 - \hat{u}(t-1)\big)\big(1 - \hat{u}(t-2)\big) \tag{15}$$

In addition, we assume a patient-dependent Markov chain likelihood (when known) / hyperprior (when unknown) for $\hat{u}(t)$, to reduce the parameter space:

$$\hat{u}(t+1) = p_{11}^{(k)} \hat{u}(t) + p_{01}^{(k)} \big(1 - \hat{u}(t)\big) \tag{16}$$

With $\hat{u}$ initialised to the steady state distribution of the Markov chain:

$$\hat{u}(t_0) = \frac{p_{01}^{(k)}}{p_{01}^{(k)} + p_{10}^{(k)}} \tag{17}$$

###### A.3.5. Measurements

We assume two measurement distributions:

- For PO-SCORAD items:

$$y_i^{(k)}(t) \sim \text{OrderedLogistic}\left(\hat{y}_i^{(k)}(t),\ (\sigma_{\mathrm{y}})_i, \boldsymbol{\delta_i}\right) \tag{18}$$

- For oSCORAD items (we exclude subjective symptoms of SCORAD, as they are the same regardless of whether the score is self-assessed or assessed by a clinician):

$$q_i^{(k)}(t) \sim \text{OrderedLogistic}\left(\hat{y}_i^{(k)}(t) + \lambda_i(t),\ (\sigma_{\mathrm{q}})_i, \boldsymbol{\delta_i}\right) \tag{19}$$

Where:

- $(\sigma_y)_i$ and $(\sigma_q)_i$ are the standard deviation of the logistic distribution.
- $\boldsymbol{\delta_i}$ is a vector of normalised distances between consecutive cutpoints.
- $\lambda_i(t)$ are the measurement biases between SCORAD and PO-SCORAD.

To calibrate PO-SCORAD measurements using SCORAD, we assume:

- SCORAD measurements are more precise than PO-SCORAD measurements (but they are not perfect)[2]:

$$\boldsymbol{\sigma}_{\mathrm{q}} = 0.5\,\boldsymbol{\sigma}_{\mathrm{y}} \tag{20}$$

- The biases $\lambda_i(t)$ decrease exponentially from $\lambda_i(t_0)$, with a characteristic time $\tau_i$ (in days):

$$\lambda_i(t) = \lambda_i(t_0)\ \exp\left(-\frac{t - t_0}{\tau_i}\right) \tag{21}$$

  As a rule of thumb, when $t - t_0 = 2\tau_i$, then the bias is equal to 14% the original bias. Considering that SCORAD measurements occur every four weeks and that the last one is at week 12, as a rule of thumb, if $\tau_i < 15$ (days), then the bias at the second measurement is almost null; and if $\tau_i > 200$ (days), we can consider the bias to be almost constant over the duration of the study.

[2]We do not let $\sigma_{\mathrm{q}}$ be inferred by the model. First, when calibrating measurements, we often assume the calibrated measurements are perfect, i.e. $\sigma_{\mathrm{q}} = 0$. As such, we do not think deciding that $\sigma_{\mathrm{q}} = 0.5\,\sigma_{\mathrm{y}}$ is more arbitrary than deciding the measurements are perfect, and is motivated by the fact clinical measurements are known to be imperfect [3]. Second, there is an identifiability issue between $\sigma_{\mathrm{q}}$ and $\sigma_{\mathrm{y}}$, as $y$ and $q$ are mathematically equivalent. Indeed, ignoring the bias term for the sake of argument, when $q$ is observed, the latent score $\hat{y}$ will be determined by $y$, $q$, and the prior that the latent score is around the previous latent score (itself informed by previous observations of $y$). As a result, the latent score will mechanically be closer to $y$ a priori, but specifying the ratio $\frac{\sigma_{\mathrm{q}}}{\sigma_{\mathrm{y}}}$ indicates to the model how “close" the latent scores should be to $q$ as opposed to $y$.

### B. Priors

Unless stated otherwise, we used weakly informative priors for our model, i.e. priors designed to rule out unreasonable parameter values without excluding any value that could make sense.

#### B.1. Power prior

##### B.1.1. Background

According to Bayes' theorem, the posterior of a (set of) parameter $\theta$ given the data $\mathcal{D}$ is:

$$p(\theta|\mathcal{D}) = \frac{p(\mathcal{D}|\theta)p(\theta)}{p(\mathcal{D})} \propto p(\mathcal{D}|\theta)p(\theta) \tag{22}$$

Where:

- $p(\mathcal{D}|\theta)$ is the likelihood.
- $p(\theta)$ is the prior.
- $p(\mathcal{D})$ is the evidence (normalisation constant).

If we have previously obtained the posterior distribution $p(\theta|\mathcal{D}_0)$ after observing the historical data $\mathcal{D}_0$, we can use this posterior as a new prior:

$$p(\theta|\mathcal{D}, \mathcal{D}_0) \propto p(\mathcal{D}|\theta)p(\theta|\mathcal{D}_0) \propto p(\mathcal{D}|\theta)p(\mathcal{D}_0|\theta)p(\theta) \tag{23}$$

If we assume that data $\mathcal{D}$ contains $N$ observations, data $\mathcal{D}_0$ contains $N_0$ observations, and the observations are independently distributed, then $p(\mathcal{D}|\theta)$ contains $N$ terms and $p(\mathcal{D}_0|\theta)$ contains $N_0$ terms. Since observations in $\mathcal{D}$ and $\mathcal{D}_0$ are weighted equally, everything else being equal, the relative contribution of data $\mathcal{D}$ to the posterior is $\frac{N}{N+N_0}$.

The power prior [4] introduces a parameter $a_0$ that weighs the contribution of the historical data $\mathcal{D}_0$:

$$p(\theta|\mathcal{D}_0) \propto p(\mathcal{D}_0|\theta)^{a_0}p(\theta) \tag{24}$$

$a_0$ quantifies how much information is borrowed from the historical data, with $a_0 = 0$ implying no borrowing and $a_0 = 1$ implying full borrowing. Put another way, as a rule of thumb, the relative contribution of the data $\mathcal{D}$ to the posterior in the presence of the power prior is $\frac{N}{N+a_0 N_0}$.

##### B.1.2. Construction of the power prior

To construct the power prior, we made several assumptions and approximations that we describe in this section.

As historical data, we used the data from an already published study investigating the role of an emollient in 337 children with AD [5]. This dataset contains 9943 patient-day observations of PO-SCORAD compared to 1136 patient-day observations in our dataset.

To ensure that the posterior is mostly determined by our dataset rather than the historical dataset, we chose $a_0 = 0.04$ (Fig. S1).

To construct the power prior, first, we used the historical data to fit the ScoradPred model, consisting of independent state-space models with ordered logistic measurement distribution and latent random walk dynamic.

Then, we exponentiate the posterior distribution rather than the likelihood to construct the power prior, resulting in initial priors being weighted by $1 + a_0$ instead of $1$ (increasing the weight results in a sharper distribution). Considering that $a_0 \ll 1$ and that the initial priors are weakly informative, this choice has little influence on the posterior, but is more convenient to implement. We also restrict the posterior distribution to the marginal distribution of the population parameters ($\boldsymbol{\sigma}_{\mathrm{l}}$, $\boldsymbol{\delta_i}$, $\boldsymbol{\sigma}_{\mathrm{y}}$, $\boldsymbol{\mu_0}$, $\boldsymbol{\sigma_0}$) rather than the full joint distribution. Finally, since distribution is represented by samples (cf. MCMC), we approximated the marginal distributions by Gaussian distributions by moment matching.

##### B.1.3. Initial prior for the measurement distributions

As explained in Section A.1, we used a weakly informative lognormal prior for the measurement noise, which translates to a 95% CI which is approximately $[0.02M, 0.40M]$, thus allowing very precise or very imprecise measurement that covers the entire range of the score.

$$(\sigma_{\mathrm{y}})_i / M_i \sim \log \mathcal{N}\big(-\log(10), \log(2)^2\big) \tag{25}$$

We used a symmetric Dirichlet prior for $\boldsymbol{\delta_i}$:

$$\boldsymbol{\delta_i} \sim \mathrm{Dirichlet}(2) \tag{26}$$

###### B.1.4. Initial prior for the latent dynamics

We used the same prior for $\boldsymbol{\sigma}_\mathrm{l}$ as for $\boldsymbol{\sigma}_\mathrm{y}$, thus allowing fast or slow transitions from a state where $y = 0$ is the most likely outcome to a state where $y = M$ is the most likely outcome:

$$(\sigma_\mathrm{l})_i / M_i \sim \log\mathcal{N}\big(-\log(10), \log(2)^2\big) \tag{27}$$

We used weakly informative priors for $\boldsymbol{\mu_0}$ and $\boldsymbol{\sigma_0}$ that translates to approximately uniform priors for $\hat{\boldsymbol{y}}_i^{(k)}(t_0)$:

$$(\mu_0)_i / M_i \sim \mathcal{N}(0.5, 0.25^2) \tag{28}$$

$$(\sigma_0)_i / M_i \sim \mathcal{N}^+(0, 0.125) \tag{29}$$

##### B.2. Correlations between severity items

We used a LKJ prior for the correlation matrix $\Omega$ of the changes in latent severity items and for the correlation matrix $\Omega_0$ of the initial latent severity items. We selected a shape parameter greater than 1, thus penalising the complexity of the model by putting more density towards the identity correlation matrix (cf. independent severity items):

$$\Omega\,,\,\Omega_0 \sim \mathrm{LKJ}(10) \tag{30}$$

##### B.3. Trend

For the trend component, we only need to define the prior for the smoothing parameter $\boldsymbol{\phi}$ and chose a prior that penalised slightly the complexity of the model, i.e. assuming more density toward a constant trend ($\boldsymbol{\phi} = \boldsymbol{0}$):

$$\phi_i \sim \mathrm{Beta}(1, 3) \tag{31}$$

##### B.4. Daily treatment usage inference

In the following, since we used the same priors for both corticosteroids and emollient cream, we drop the subscript $j$ indexing treatment out of concision.

We used the same priors for the Markov chain parameters $p_{10}^{(k)} = 1 - p_{11}^{(k)}$ and $p_{01}^{(k)}$. We defined a hierarchical prior for $p_{10}^{(k)}$ and $p_{01}^{(k)}$, and chose weakly informative priors for the population mean

($\mu_{10}$, $\mu_{01}$) and standard deviation ($\sigma_{10}$, $\sigma_{01}$) parameters, that translate to approximately uniform distribution for $p_{10}^{(k)}$ and $p_{01}^{(k)}$:

$$
\begin{aligned}
p_{10}^{(k)} &\sim \operatorname{logit} \mathcal{N}(\mu_{10}, \sigma_{10}^2) && (32)\\
p_{01}^{(k)} &\sim \operatorname{logit} \mathcal{N}(\mu_{01}, \sigma_{01}^2) && (33)\\
\mu_{10}\,,\ \mu_{01} &\sim \mathcal{N}(0, 1) && (34)\\
\sigma_{10}\,,\ \sigma_{01} &\sim \mathcal{N}^{+}(0, 1.5) && (35)
\end{aligned}
$$

#### B.5. Treatment effects

We used the same weakly informative priors for both treatment and all severity items, assuming that treatment effects are approximately within $\pm$ 20% the range of the score:

$$\Theta_{i,j}/M_i \sim \mathcal{N}(0, 0.1^2) \tag{36}$$

#### B.6. Calibration

We assumed the prior for initial bias $\lambda_i(t_0)$ to be within $\pm$ 20% the range of the severity item:

$$\lambda_i(t_0)/M_i \sim \mathcal{N}(0, 0.1^2) \tag{37}$$

We assumed a lognormal prior for the characteristic learning time $\tau_i$, such as most of its mass is between 1 day and $\approx$ 250 days, which allows very fast learning (bias null for at the second SCORAD measurement) or no learning (constant bias for the duration of the study):

$$\tau_i \sim \log \mathcal{N}\Big(1.2 \log(10), \big(0.6 \log(10)\big)^2\Big) \tag{38}$$

### C. Treatment recommendations

#### C.1. Utility function

We consider two binary actions (using or not using topical corticosteroids $TC$ or emollient cream $EC$), not mutually exclusive, that we denote with $a = \{TC, EC\} \in \{0, 1\}^2$.

We define the utility function as a function of the action $a$ and the predicted PO-SCORAD for that action $\hat{y}(a)$ by:

$$U\big(\hat{y}(a), a\big) = -\big(\hat{y} + cost(a)\big) \tag{39}$$

Where $cost(a)$ is the “perceived" cost of action $a$, that we define by:

$$cost(a) = cost_{TC} \times TC + cost_{EC} \times EC + cost_{both} \times TC \times EC \tag{40}$$

Where:

- $cost_{TC}$ and $cost_{EC}$ are the costs of using topical corticosteroids and emollient cream, respectively.
- $cost_{both}$ is the additional cost when using the two treatments.

It is worth noting that:

- The cost of using “no treatment" is set to 0, without loss of generality.
- The $cost(a)$ is in the same unit as $\hat{y}$ (i.e. PO-SCORAD), and can be interpreted as the minimum improvement in $y$ the patient would require to use treatment.

#### C.2. Objective function

We chose to maximise the following objective function, which includes a risk-sensitive criterion:

$$E\Big(U\big(\hat{y}(a), a\big)\Big) - z\sqrt{V\Big(U\big(\hat{y}(a), a\big)\Big)} \tag{41}$$

Where $z$ quantifies the tolerance to risk (uncertainty):

- $z$ can be interpreted as a z-score, assuming the utility is normally distributed. For example, if $z = 1.96$, the objective function is the lower bound of the 95% CI of the utility, i.e. the 2.5% quantile.

- $z > 0$ corresponds to a patient that is risk-averse (penalising uncertainty) or pessimistic (maximise the worst case).
- $z < 0$ corresponds to a patient that is risk-seeking (welcoming uncertainty) or optimistic (maximise the best case). $z < 0$ encourages the exploration of new treatments with uncertain effects.
- $z = 0$ corresponds to a patient that is risk-neutral.

#### C.3. Decision profiles

In our situation, the decision profile of a patient is fully determined by its sensitivity to risk $z$ and its "perceived" cost of using treatment, parametrised by $cost_{TC}$, $cost_{EC}$ and $cost_{both}$.

We defined three risk profiles (risk-averse, risk-neutral and risk-seeking) and three cost profiles (no cost, normal cost, high cost) for a total of nine decision profiles (Table S1). The cost profiles assume the same costs for topical corticosteroids and emollient cream.

Even though we believe these decision profiles to be relevant, they are only illustrative and do not claim to represent the preferences of an actual patient. In particular, the decision parameters may change to reflect changes in patients' preferences. For example, $z$ could be negative at the beginning of a trial to encourage the exploration of new treatments and be increased gradually to exploit a treatment that has been proven to be effective.

Table S1: Decision profiles for treatment recommendations

| **Decision profile** | **Cost profile** | **Risk Profile** | **Treatment costs** | | | **Risk tolerance** |
|---|---|---|---|---|---|---|
| | | | $cost_{TC}$ | $cost_{EC}$ | $cost_{both}$ | $z$ |
| 1 | No cost | Risk averse | 0 | 0 | 0 | 1.5 |
| 2 | No cost | Risk seeking | 0 | 0 | 0 | 0 |
| 3 | No cost | Risk neutral | 0 | 0 | 0 | -1.5 |
| 4 | Normal cost | Risk averse | 0.5 | 0.5 | 0 | 1.5 |
| 5 | Normal cost | Risk seeking | 0.5 | 0.5 | 0 | 0 |
| 6 | Normal cost | Risk neutral | 0.5 | 0.5 | 0 | -1.5 |
| 7 | High cost | Risk averse | 3 | 3 | 3 | 1.5 |
| 8 | High cost | Risk seeking | 3 | 3 | 3 | 0 |
| 9 | High cost | Risk neutral | 3 | 3 | 3 | -1.5 |

### References

[1] G. Hurault, J. F. Stalder, S. Mery, *et al.*, "EczemaPred: A computational framework for personalised prediction of eczema severity dynamics," *Clinical and Translational Allergy*, vol. 12, no. 3, e12140, Mar. 2022, ISSN: 2045-7022. DOI: 10.1002/clt2.12140 (cit. on p. 2).

[2] R. J. Hyndman and G. Athanasopoulos, *Forecasting : principles and practice.* OTexts: Melbourne, Australia, 2018, p. 291, ISBN: 9780987507105 (cit. on p. 5).

[3] J. Schmitt, S. Langan, S. Deckert, *et al.*, "Assessment of clinical signs of atopic dermatitis: a systematic review and recommendation.," *The Journal of allergy and clinical immunology*, vol. 132, no. 6, pp. 1337–47, Dec. 2013, ISSN: 1097-6825. DOI: 10.1016/j.jaci.2013.07.008 (cit. on p. 7).

[4] J. G. Ibrahim, M. H. Chen, Y. Gwon, and F. Chen, "The power prior: Theory and applications," *Statistics in Medicine*, vol. 34, no. 28, pp. 3724–3749, 2015, ISSN: 10970258. DOI: 10.1002/sim.6728 (cit. on p. 8).

[5] G. S. Tiplica, F. Boralevi, P. Konno, *et al.*, "The regular use of an emollient improves symptoms of atopic dermatitis in children: a randomized controlled study," *Journal of the European Academy of Dermatology and Venereology*, vol. 32, no. 7, pp. 1180–1187, Jul. 2018, ISSN: 14683083. DOI: 10.1111/jdv.14849 (cit. on p. 9).

### Supplementary Figures

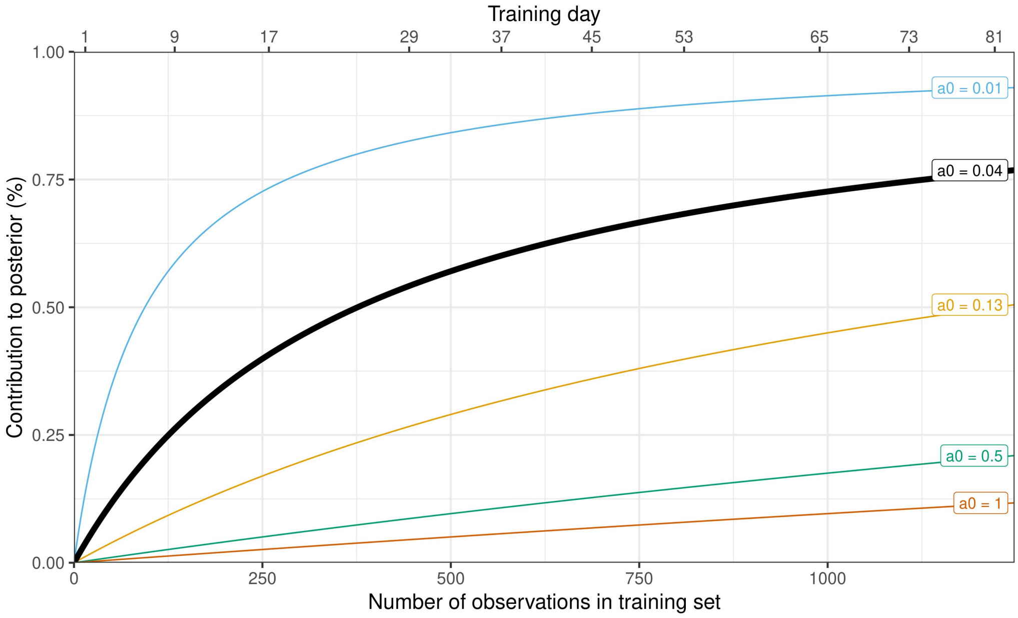


Figure S1: Approximate contribution of our dataset to the posterior distribution as a function of the number of observations in the training set, for different values of the power prior's discounting parameter $a_0$. We use $a_0 = 0.04$, which can be interpreted as if the model was pre-trained with $a_0 \times N_{\text{historical}} = 0.04 \times 9943 \approx 400$ observations.

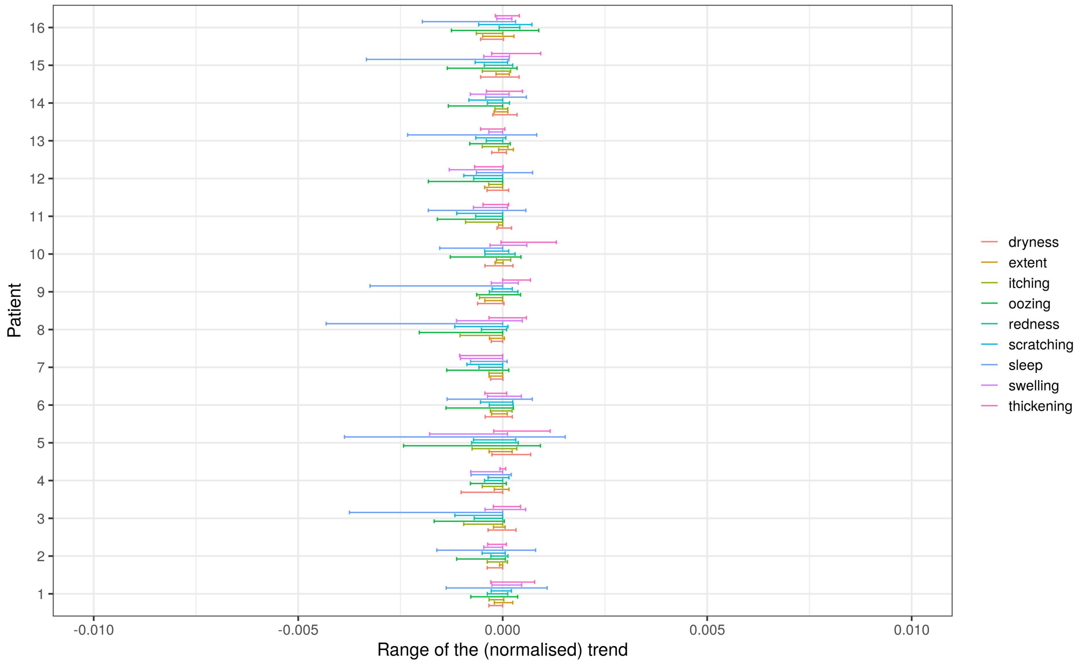


Figure S2: Minimum and maximum of the expected trend component, for each patient and each severity item. The estimates are normalised by the range of the score. For example, if the maximum is 0.01, for extent (defined in $[0, 100]$) this would mean that the maximum expected trend is $0.01 \times 100 = 1$. We can consider that the trend is always zero as the order of magnitude of the amplitude of the trend is around 0.001.

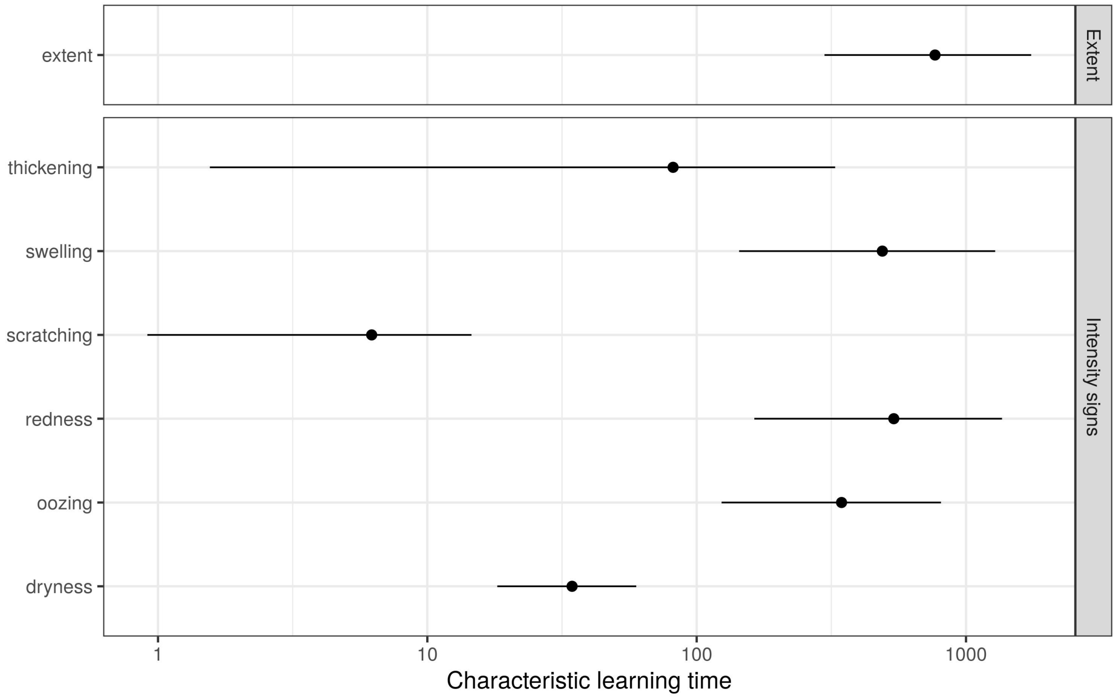


Figure S3: Mean and 90% credible interval of the characteristic learning time $\tau$ of the calibration process (in days). Since SCORAD is measured every four weeks, estimates less than $\approx 15$ days would translate to a bias of 0 for the second measurement. Any estimates greater than $\approx 200$ days (longer than the study follow-up) can be interpreted as a constant bias (no learning).

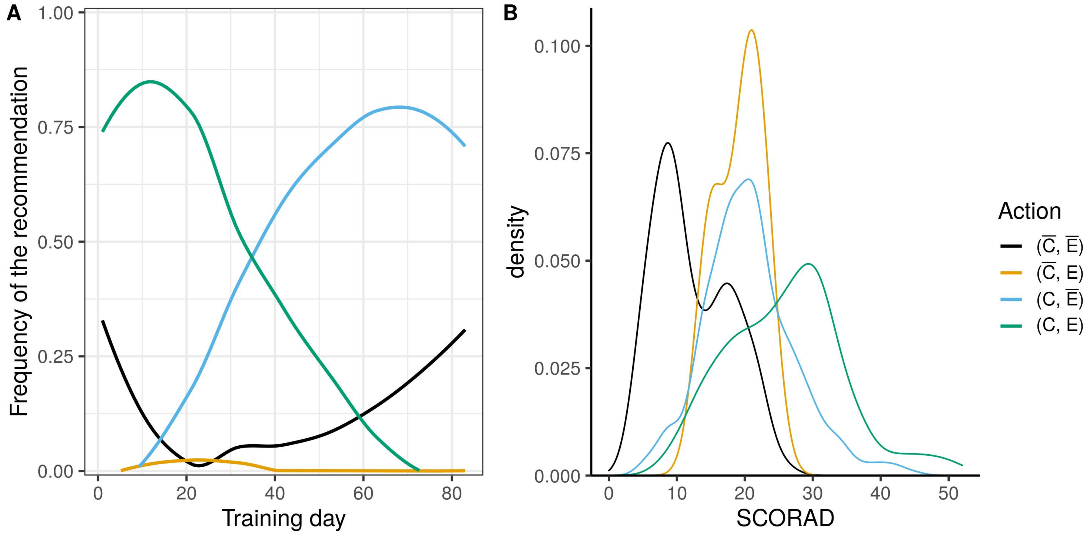


Figure S4: Analysis of treatment recommendations for a risk-neutral patient and a “normal" perceived cost of treatments. $C$ and $\bar{C}$ correspond to the action of using and not using corticosteroids, respectively; $E$ and $\bar{E}$ correspond to the action of using and not using emollients, respectively. A) Frequency of recommended actions as a function of training day, smoothed by LOWESS. B) Distribution of SCORAD at the time of the recommendation, for each action. Overlapping distributions suggests that multiple actions could be recommended for a same SCORAD.

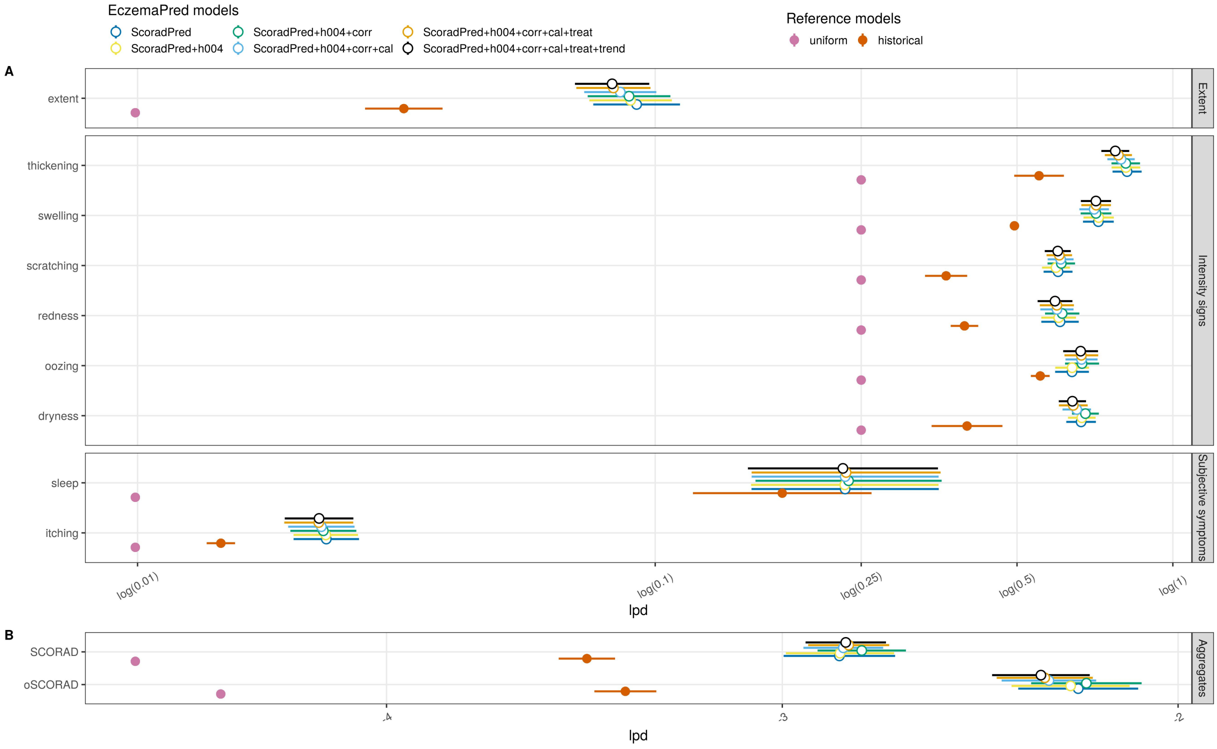


Figure S5: Predictive performance (lpd) estimates (mean ± SE) for four-days-ahead predictions after training the model with 65 days of data (79% of the data). "ScoradPred" corresponds to the base model with independent state-space models for each severity item, no power prior, calibration, treatment effects or trend. The suffixes indicate the additions made to the base model, where "h004" corresponds to the power prior with $a_0 = 0.04$, "corr" to the correlation between severity items, "cal" to the calibration data, "treat" to treatment effects and "trend" to the trend component. A) Severity items predictive performance. B) (o)SCORAD predictive performance.
